# Illness characteristics of COVID-19 in children infected with the SARS-CoV-2 Delta variant

**DOI:** 10.1101/2021.10.06.21264467

**Authors:** Erika Molteni, Carole H. Sudre, Liane S. Canas, Sunil S. Bhopal, Robert C. Hughes, Liyuan Chen, Jie Deng, Benjamin Murray, Eric Kerfoot, Michela Antonelli, Mark Graham, Kerstin Kläser, Anna May, Christina Hu, Joan Capdevila Pujol, Jonathan Wolf, Alexander Hammers, Tim D. Spector, Sebastien Ourselin, Marc Modat, Claire J. Steves, Michael Absoud, Emma L. Duncan

## Abstract

**Background:** The Delta (B.1.617.2) SARS-CoV-2 variant became the predominant UK circulating strain in May 2021. Whether COVID-19 from Delta infection differs to infection with other variants in children is unknown.

**Methods:** Through the prospective COVID Symptom Study, 109,626 UK school-aged children were proxy-reported between December 28, 2020 and July 8, 2021. We selected all symptomatic children who tested positive for SARS-CoV-2 and were proxy-reported at least weekly, within two timeframes: December 28, 2020 to May 6, 2021 (Alpha (B.1.1.7) the main UK circulating variant); and May 26 to July 8, 2021 (Delta the main UK circulating variant). We assessed illness profiles (symptom prevalence, duration, and burden), hospital presentation, and presence of long (≥28 day) illness; and calculated odds ratios for symptoms presenting within the first 28 days of illness.

**Findings:** 694 (276 younger [5-11 years], 418 older [12-17 years]) symptomatic children tested positive for SARS-CoV-2 with Alpha infection and 706 (227 younger and 479 older) children with Delta infection. Median illness duration was short with either variant (overall cohort: 5 days (IQR 2–9.75) with Alpha, 5 days (IQR 2-9) with Delta). The seven most prevalent symptoms were common to both variants. Symptom burden over the first 28 days was slightly greater with Delta compared with Alpha infection (in younger children, 3 (IQR 2–5) with Alpha, 4 (IQR 2–7) with Delta; in older children 5 (IQR 3–8) with Alpha and 6 (IQR 3–9) with Delta infection in older children). The odds of several symptoms were higher with Delta than Alpha infection, including headache and fever. Few children presented to hospital, and long illness duration was uncommon, with either variant.

**Interpretation:** COVID-19 in UK school-aged children due to SARS-CoV-2 Delta strain B.1.617.2 resembles illness due to the Alpha variant B.1.1.7., with short duration and similar symptom burden.

**Funding:** ZOE Limited, UK Government Department of Health and Social Care, Wellcome Trust, UK Engineering and Physical Sciences Research Council, UK Research and Innovation London Medical Imaging & Artificial Intelligence Centre for Value Based Healthcare, UK National Institute for Health Research, UK Medical Research Council, British Heart Foundation and Alzheimer’s Society.

**Ethics:** Ethics approval was granted by KCL Ethics Committee (reference LRS-19/20-18210).

**Research in context:** *Evidence before this study:* To identify existing evidence for differences in COVID-19 due to infection with Alpha (B.1.1.7) or Delta (B.1.617.2) SARS-CoV-2 variants, we searched PubMed for peer-reviewed articles and medRxiv for preprint publications between March 1, and September 17, 2021 using keywords (“SARS-CoV-2” OR “COVID-19”) AND (children OR p?ediatric*) AND (“delta variant” OR “B.1.617.2”). We did not restrict our search by language. Of twenty published articles identified in PubMed, we found one case study describing disease presentation associated with Delta variant infection in a child. Another study examining the increase in hospitalization rates of paediatric cases in USA from August 1, 2020 to August 27, 2021 stated that “It is not known whether the B.1.617.2 (Delta) variant […] causes different clinical outcomes in children and adolescents compared with variants that circulated earlier.” Four studies reported cases of transmission of the Delta variant in school and community contexts; and two discussed screening testing in school-aged children (thus not directly relevant to the research question here). Remaining papers did not target paediatric age specifically. We found no studies investigating differences in COVID-19 presentation (e.g., duration, burden, individual symptoms) in school-aged children either in the UK or world-wide.

*Added value of this study:* We describe and compare illness profiles in symptomatic UK school-aged children (aged 5–17 years) with COVID-19 when either Alpha or Delta strains were the predominant circulating SARS-CoV-2 variant. Our data, collected through one of the largest UK citizen science epidemiological initiatives, show that symptom profile and illness duration of COVID-19 are broadly similar between the strains. Although there were slightly more symptoms with Delta than with Alpha, particularly in older children, this was offset by similar symptom duration (whether considered for symptoms individually or for illness overall). Our study adds quantitative information to the debate on whether there are meaningful clinical differences in COVID-19 due to Alpha vs. Delta variants; and contributes to the discussion regarding rationale for vaccinating children (particularly younger children) against SARS-CoV-2.

*Implications of all the available evidence:* Our data confirm that COVID-19 in UK school-aged children is usually of short duration and similar symptom burden, whether due to Delta or Alpha. Our data contribute to epidemiological surveillance from the wider UK population, and we capture common and generally mild paediatric presentations of COVID-19 that might be missed using clinician-based surveillance alone. Our data will also be useful for the vaccination debate.

## Introduction

Viruses acquire genetic change over time,^1^ which can alter transmissibility,^2^ virulence,^3^ clinical presentation,^4,5^ and/or effectiveness of vaccination and other health measures.^6^ Correspondingly, multiple SARS-CoV-2 variants have emerged across the pandemic, with four variants of concern and ten variants under investigation circulating internationally at the time of writing (https://www.who.int/en/activities/tracking-SARS-CoV-2-variants/). The Alpha variant (B.1.1.7) initially emerged in the UK in September 2020 (https://www.who.int/en/activities/tracking-SARS-CoV-2-variants/), and was the predominant UK strain from November 2020 until early May 2021 (https://assets.publishing.service.gov.uk/government/uploads/system/uploads/attachment_data/file/959438/Technical_Briefing_VOC_SH_NJL2_SH2.pdf; https://assets.publishing.service.gov.uk/government/uploads/system/uploads/attachment_data/file/1014926/Technical_Briefing_22_21_09_02.pdf). The Delta variant (B.1.617.2) was initially identified in India in October 2020, with the Delta lineage designated a “variant of concern” by WHO on May 11, 2021 (https://www.who.int/en/activities/tracking-SARS-CoV-2-variants/). By the end of May 2021, the Delta variant was the predominant (>75%) circulating variant in the UK, reaching >99% in the week commencing on June 27, 2021 (Supplementary Table 1: https://assets.publishing.service.gov.uk/government/uploads/system/uploads/attachment_data/file/1018547/Technical_Briefing_23_21_09_16.pdf). The Delta variant appears more transmissible than wild-type or other emergent variants to date, with transmissibility approximately 60% higher than the Alpha variant.^7,8^ There is some evidence that COVID-19 due to infection with the Delta variant may be more severe than previous strains, with increased requirements for hospitalization and respiratory support;^9,10^ and currently available vaccines may be modestly less efficacious against infection with the Delta variant than preceding variants, although still very effective against severe illness and death.^11^

In adults, comparisons of COVID-19 caused by the Delta variant vs. other variants is particularly complicated by vaccination roll-out and altered clinical presentation in individuals infected post-vaccination.^12^ However, children in the UK were not offered vaccination until after Delta became the main circulating variant. At-risk minors (i.e., with severe neuro-disabilities, Down’s syndrome, immunosuppression, multiple or severe learning disabilities) aged ≥12 years, and those co-living with immunosuppressed individuals, were offered vaccination after July 19, 2021; a single dose was offered to children aged 16-17 years after August 4, 2021 and to children aged ≥12 years after September 13, 2021 (date of announcement; https://www.gov.uk/government/news/jcvi-issues-advice-on-covid-19-vaccination-of-children-and-young-people).

We previously described illness characteristics in UK school-aged children with COVID-19 who tested positive between September 1, 2020 and January 24, 2021,^13^ during which time the predominant circulating variants were wild-type and the Alpha variant (https://www.gov.uk/government/publications/covid-19-variants-genomically-confirmed-case-numbers). Here, we describe and compare illness profiles in symptomatic UK school-aged children with SARS-CoV-2 infection presenting during two timeframes: December 28, 2020 (during the peak of the winter wave in the UK) to May 6, 2021, when the Alpha variant was the main UK circulating variant; and May 26, 2021 to July 8, 2021, when Delta was the predominant variant (https://www.gov.uk/government/publications/covid-19-variants-genomically-confirmed-case-numbers).

## Methods

This study used data from COVID Symptom Study, using the ZOE COVID Study App (details previously published^13^). Briefly, this prospective study collects self-reported data from adult participants through a mobile application (app), including direct questions about specific symptoms (Supplementary Table 2), free text symptom reporting, SARS-CoV-2 testing, vaccination, and health care access, as well as demographic and co-morbidity data. Adult contributors can proxy-report for children; however, data cannot be linked between contributor and proxy-reported individual. Children aged 16-17 can also self-report. Questions about symptoms and comorbidities were largely informed by adult data,^14^ and do not include some common paediatric co-morbidities (e.g., neurological or neuro-disability disorders). Ethics approval for this study was granted by the KCL Ethics Committee (LRS-19/20-18210). Upon registration through the app, all participants provided consent for their data to be used for COVID-19 research. Governance was specifically granted to enable use of proxy-reported data including data from minors.

Data from all proxy-reported UK school-aged children (5–17 years) with a positive test between December 28, 2020 to July 8, 2021 (i.e., prior to vaccination approval for any child in the UK) was selected. As previously,^15^ children were considered to have COVID-19 if proxy-reported with relevant symptoms between two weeks before and one week after SARS-CoV-2 infection confirmation (either PCR or LFAT). Data were included for children proxy-reported at least once weekly, from first symptom report until healthy or until proxy-reporting ceased, noting our previous study showed proxy-reporting in this cohort is assiduous (continued logging until healthy in >90% of children).^13^

Illness characteristics were assessed for children logging a positive test within two timeframes, according to whether Alpha or Delta was the predominant circulating UK SARS-CoV-2 variant at the time (Supplementary Table 1). The first was from December 28, 2020 to May 6, 2021 (noting December 29, 2020 was the peak positive specimen date for 2020-2021 UK winter wave of the pandemic, https://assets.publishing.service.gov.uk/government/uploads/system/uploads/attachment_data/file/975754/Variants_of_Concern_Technical_Briefing_8_Data_England.xlsx). The second was from May 26, 2021 to July 8, 2021 (https://assets.publishing.service.gov.uk/government/uploads/system/uploads/attachment_data/file/959359/Variant_of_Concern_VOC_202012_01_Technical_Briefing_4. pdf AND https://assets.publishing.service.gov.uk/government/uploads/system/uploads/attachment_data/file/1018476/Variants_of_Concern_Technical_Briefing_23_Data_Englan_d.xlsx AND https://assets.publishing.service.gov.uk/government/uploads/system/uploads/attachment_data/file/1018547/Technical_Briefing_23_21_09_16.pdf). Individual test results regarding variant strain were not available; thus, infection presenting in the first timeframe was assumed to be due to the Alpha variant and in the second to the Delta variant; terminology herein reflects these assumptions. Data were analysed overall and within two age groups: younger children aged 5–11 years (UK primary school-aged children) and older children aged 12–17 years (UK secondary school-aged children, noting that children can leave school after age 16 years).

Illness duration was calculated from first symptom (having been previously asymptomatic) until recovery (return to asymptomatic or, if proxy-reporting ceased before becoming asymptomatic, final proxy report). Individuals proxy-reported as asymptomatic but subsequently re-reported with symptoms within one week of their last symptomatic report were considered unwell from initial presentation (i.e., having relapsing or remitting illness) and illness duration calculated accordingly. Individual symptom prevalence was assessed, according to duration between first and last report for that symptom. Participant selection was determined by date of positive SARS-CoV-2 testing; however, symptoms were considered over the entire illness duration, which could extend outside the test date boundaries to a maximum of two weeks before (by virtue of definition of COVID-19 for this study) and four weeks after the timeframe boundaries (by virtue of data censoring). Data were censored on August 5, 2021, 28 days after the last date of inclusion for children whose illness commenced during the Delta period; thus, duration of individual symptoms and of overall illness for children who were ill for >28 days could not be robustly determined for every child; instead, we report the number of children who were still unwell at 28 days for both timeframes, and exclude them from descriptive statistics regarding symptom and illness duration otherwise. Saliently, our previous study of COVID-19 in children did not demonstrate new symptoms appearing later than 28 days after a positive test;^13^ similarly, we scrutinised data from children with long illness duration for new symptoms presenting after Day 28 for the current study. Odds ratios were calculated to compare symptom presentation with Delta vs. Alpha infection, considering all symptoms presenting within 28 days. Symptom burden (number of symptoms) was calculated over two timeframes: within first week (<7 days) and over entire course of illness (considered to 28 days). Symptom burden was considered for all symptoms; and with respect to the seven most common symptoms for each variant, which seven symptoms proved common to both cohorts (considered across the entire age range). Children presenting to hospital (comprising presentation to the emergency department or admission to hospital) for each timeframe were also counted.

As a proxy for other circulating viruses and possible co-infection influencing illness profile, we considered symptoms in children who tested negative for SARS-CoV-2 within the same timeframes; and provide context with Public Health England data for circulating viruses other than COVID-19 (Public Health England DataMart estimates based on sentinel laboratory surveillance; https://assets.publishing.service.gov.uk/government/uploads/system/uploads/attachment_data/file/1016276/Weekly_Flu_and_COVID-19_report_w36.pdf figure 16).

### Statistical analyses

Data are presented as descriptive statistics. Tests for differences on counts and raw prevalence were not conducted, given the unmatched nature of the cohorts and the many differences between the two timeframes (including school opening, social distancing regulations, and changing seasons). Where robust comparisons were possible (e.g., demographic data), Wilcoxon signed-rank testing, two tailed χ^2^-tests, or Fisher’s exact tests were used. Odds ratios were calculated using logistic regression for all symptoms, unless having null prevalence, with age and gender as covariates. Body mass index was not used as its robustness and utility are debated for paediatric cohorts. False Discovery Rate (FDR) correction was applied to multiple tests on non-independent data to adjust the level of statistical significance.^16^ All analyses were performed in Python version 3.7.

## Results

Overall, 109,626 UK children aged 5-17 years (50,832 younger and 58,794 older children) were proxy-reported between December 28, 2020 and July 8, 2021. Of these, 60,050 (20,054 younger and 39,996 older children) were tested for SARS-CoV-2 (Figure 1). Considering symptomatic children testing positive for SARS-CoV-2, with sufficient proxy-logging for illness profiling: 694 children (276 younger, 418 older children) tested positive between December 28, 2020 and May 6, 2021, assumed to be with Alpha variant; 706 children (227 younger, 479 older children) tested positive between May 26, 2021 and July 8, 2021 (with symptom onset on or before 8 July in all children), assumed to be with Delta variant (Table 1).

**Figure 1.**
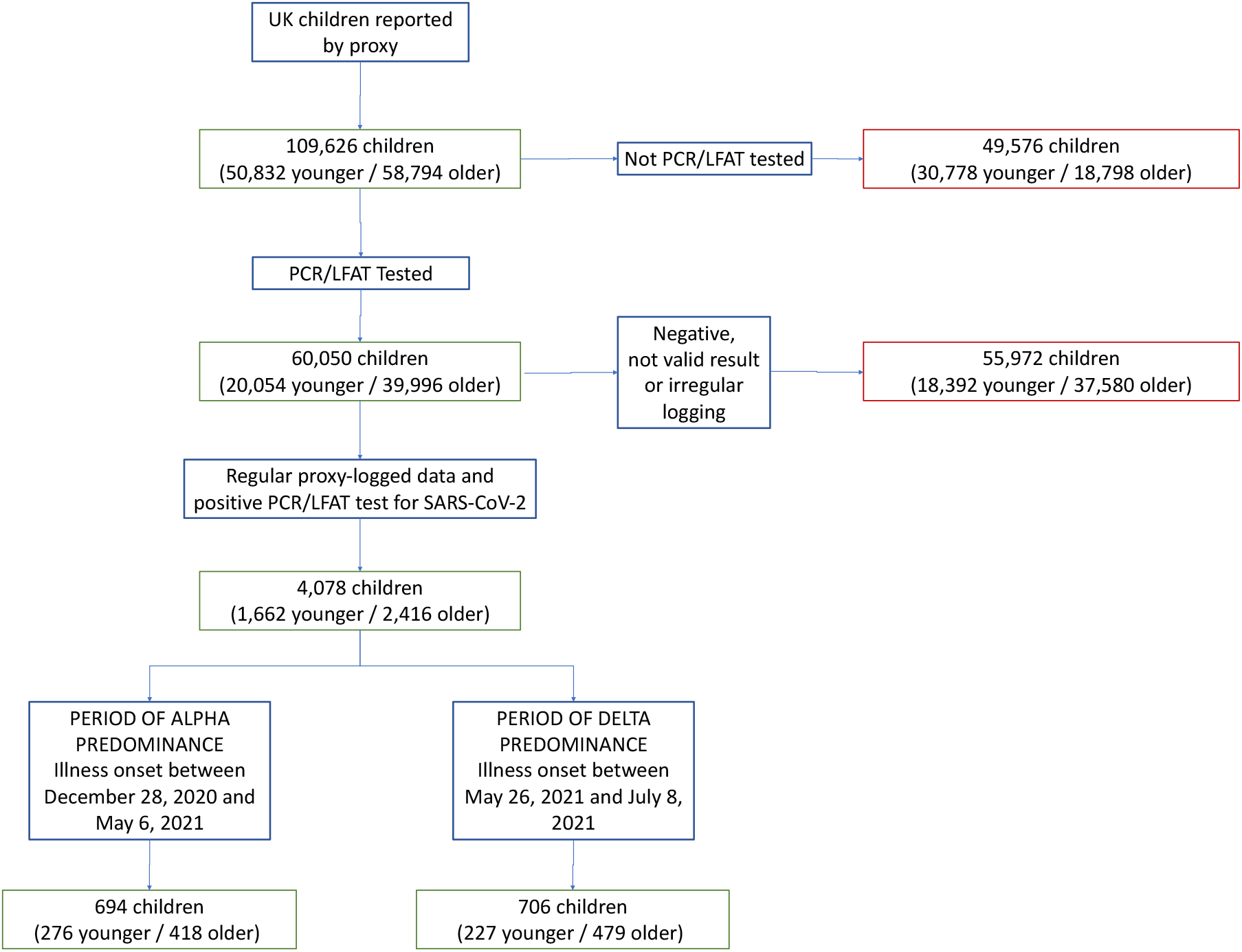
Study flowchart of inclusion and exclusion criteria. Overall number for the entire cohort of children is given first. Younger children = aged 5–11 years (UK primary school-aged children). Older children = aged 12–17 years (UK secondary school-aged children). Not valid result = test result proxy-reported as “failed test” or “still waiting”. Irregular logging = proxy-reporting with intervals of more than 7 days between proxy-reports during illness.

**Median illness duration** with COVID-19 in the cohort overall was 5 days (IQR 2– 9.75) with Alpha and 5 days (IQR 2-9) with Delta infection. Illness duration in younger children was four (IQR 2–8) days with Alpha and four (IQR 2–7.5) days with Delta infection, and in older children six (IQR 3–10) days with Alpha and six (IQR 3– 10) days with Delta infection. Although illness duration appeared slightly shorter in younger children than older children within each variant’s time period, this was not significant (younger children vs. older children during the Alpha period Z= 0.53, p =0.594, and during the Delta period Z= 0.74, p=0.458).

**Individual symptom prevalence in children with Alpha or Delta infection are** presented in Figure 2 and Supplementary Table 3. In younger children the commonest symptoms were the same with either variant: rhinorrhoea (“runny nose”) (130 [47·1%] of 276), headache (110 [39·9%] of 276), and fatigue (107 [38·8%] of 276) with Alpha infection; and headache (138 [60·8%] of 227), rhinorrhoea (122 [53·7%] of 227), and fatigue (111 [48·9%] of 227) with Delta infection. In older children with Alpha infection, the three commonest symptoms were again headache (257 [61·5%] of 418), fatigue (238 [56.9%] of 418), and rhinorrhoea (198 [47.4%] of 418); while for Delta they were headache (353 [73.7%] of 479), sore throat (290 [60.5%] of 479) and fatigue (288 [60.1%] of 479). The top seven symptoms for each cohort were common to both variants: headache, fatigue, fever, dysosmia (encompassing both anosmia and dysosmia), sneezing, rhinorrhoea, and sore throat, although there were some differences in prevalence order (Figure 2).

**Figure 2.**
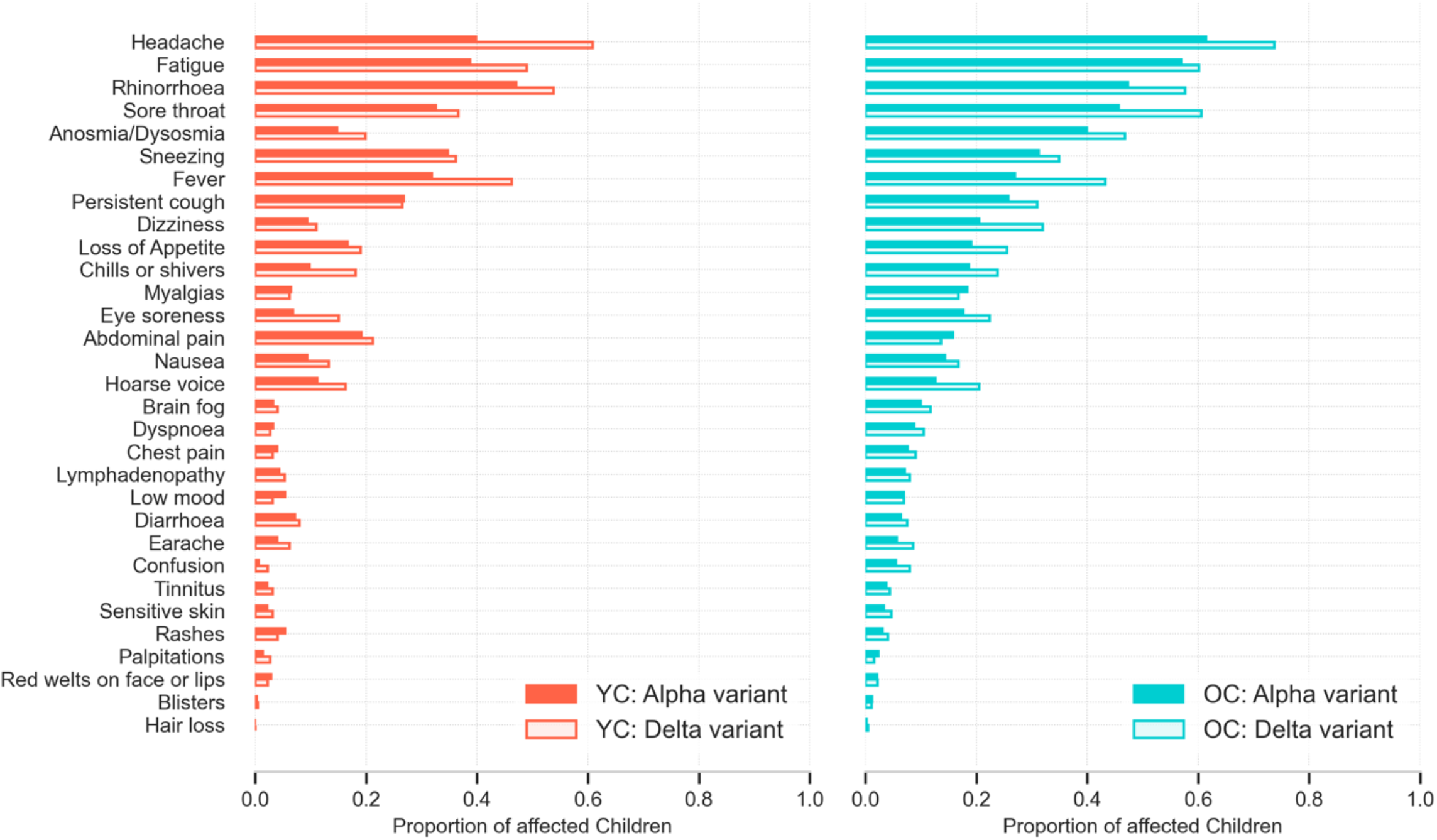
Prevalence of symptoms reported over the course of illness (to 28 days) in younger (5–11 years) and older (12–17 years) children with COVID-19 during periods of SARS-CoV-2 Alpha or Delta variant predominance.

**Odds ratios** were computed for any symptom presenting by day 28, comparing Delta to Alpha infection (Figure 3, Supplementary Table 4). Significance was determined after FDR correction, thus some odds ratios that do not cross the unit value were not significant. Considered overall, several symptoms were more common with Delta than with Alpha infection: headache, rhinorrhoea, sore throat dysosmia, fever, dizziness, “chills or shivers”, eye soreness, and hoarse voice. Considered within age groups, six symptoms were more common with Delta compared with Alpha infection in older children, and three in younger children.

**Figure 3.**
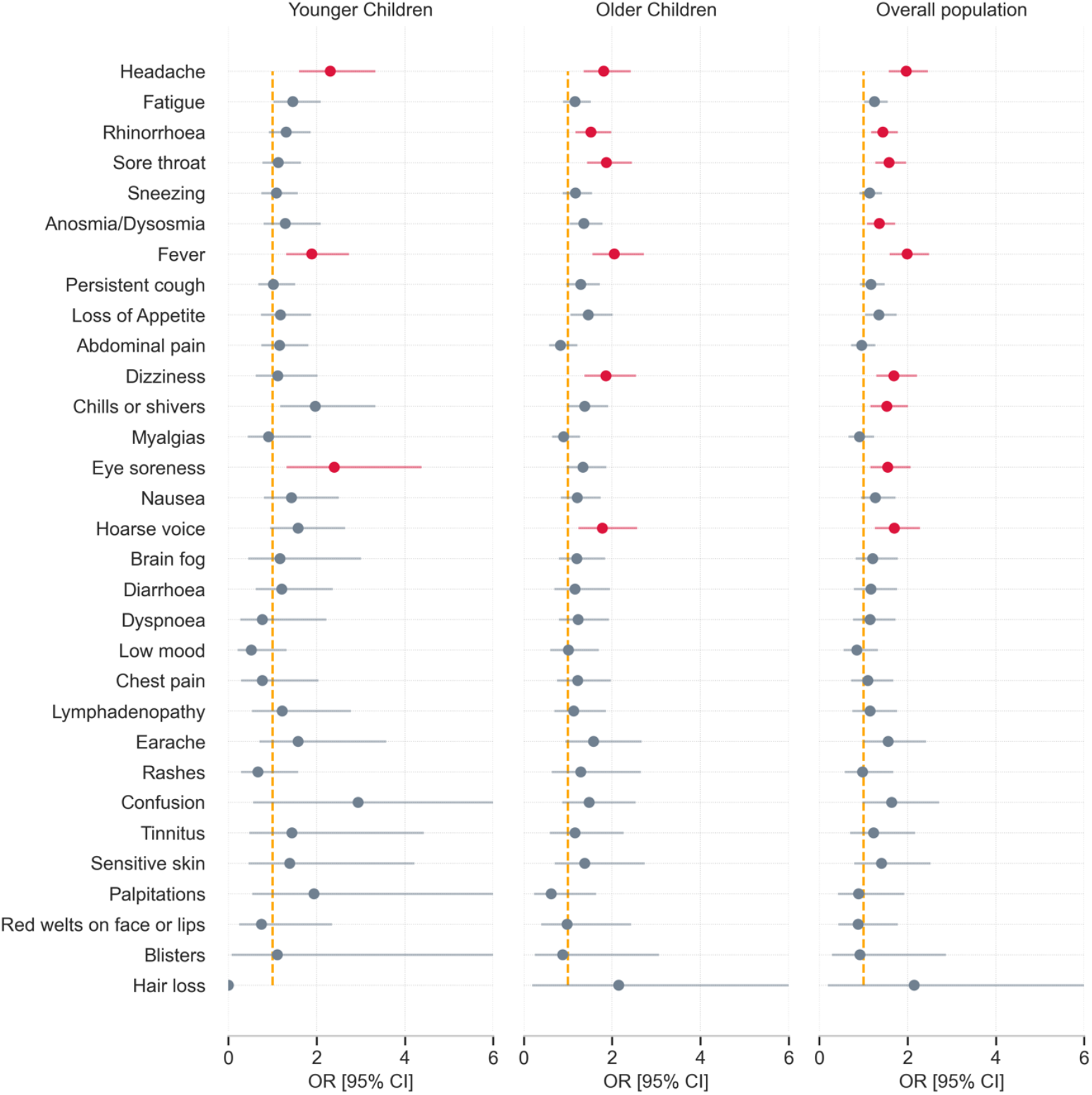
Odds ratios for a symptom presenting within the first 28 days of illness in children with COVID-19 during periods of SARS-CoV-2 Delta vs. Alpha variant predominance. Results for younger (5-11 years) and older (12-17 years) children, and for the cohort overall, comparing symptom prevalence during Delta with prevalence during Alpha infection, and adjusted for age and sex. Results with statistical significance at False Discovery Rate test (α=0·05) are in red.

**Median symptom burden, considered over the entire illness**, was 3 (IQR 2–5) with Alpha, and 4 (IQR 2–7) with Delta infection in younger children; and 5 (IQR 3–8) symptoms with Alpha and 6 (IQR 3–9) with Delta infection in older children. **Considering symptoms experienced in the first week of illness only** (noting here that median illness duration was less than a week in both age groups with either variant), median symptom burden was 3 (IQR 2–5) with Alpha and 4 (IQR 2–6) with Delta infection in younger children; and 5 (IQR 3–7) with Alpha and 6 (IQR 3–9) with Delta infection in older children (Table 1). **Considering the seven commonest symptoms** (as above, common to both variants), the median burden was 2 (IQR 1-3) with Alpha and 3 (IQR 2-4) with Delta infection in younger children, and 3 (IQR 2-4) with Alpha and 4 (IQR 2-5) with Delta in older children.

**Table 1.**
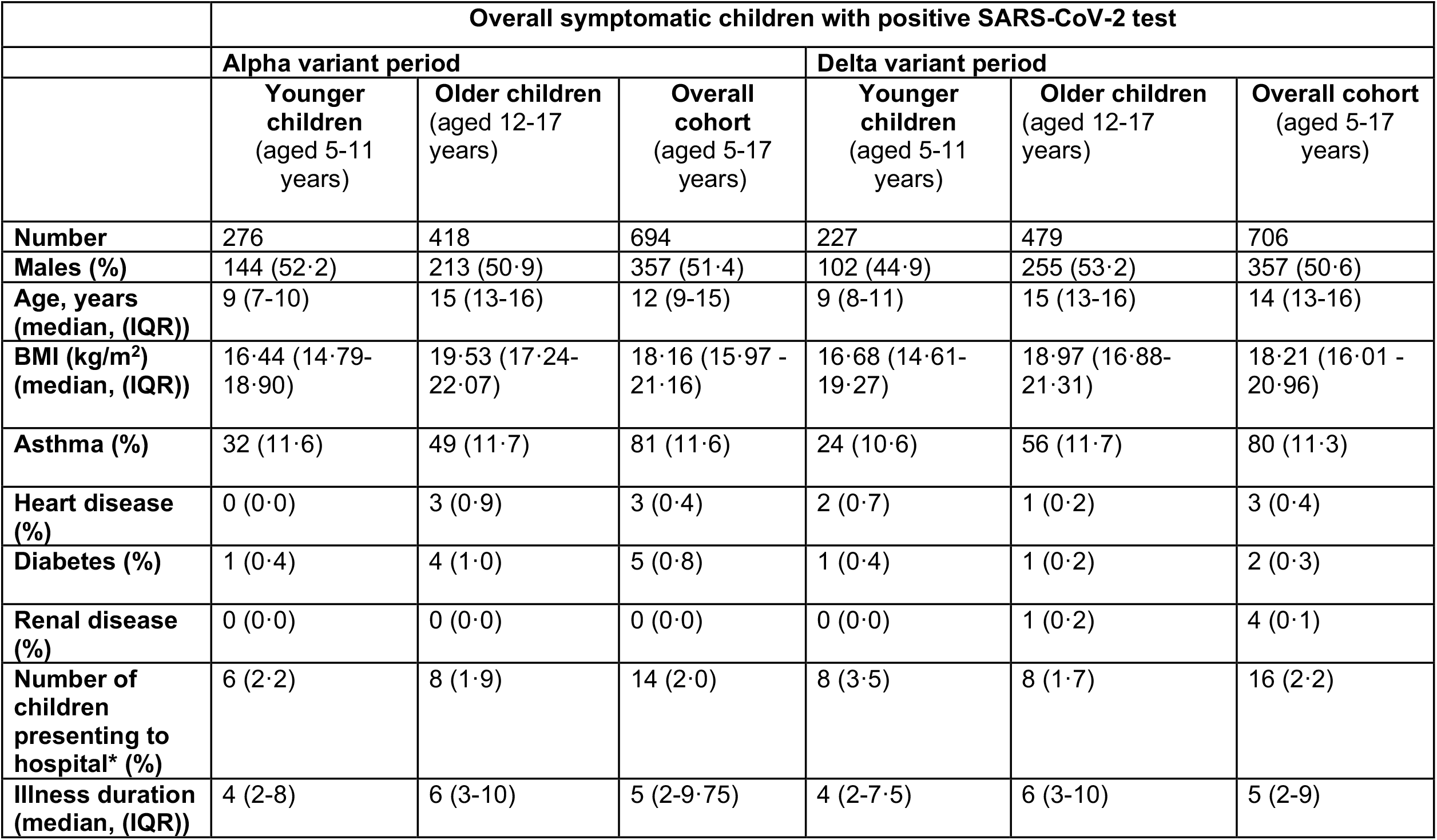
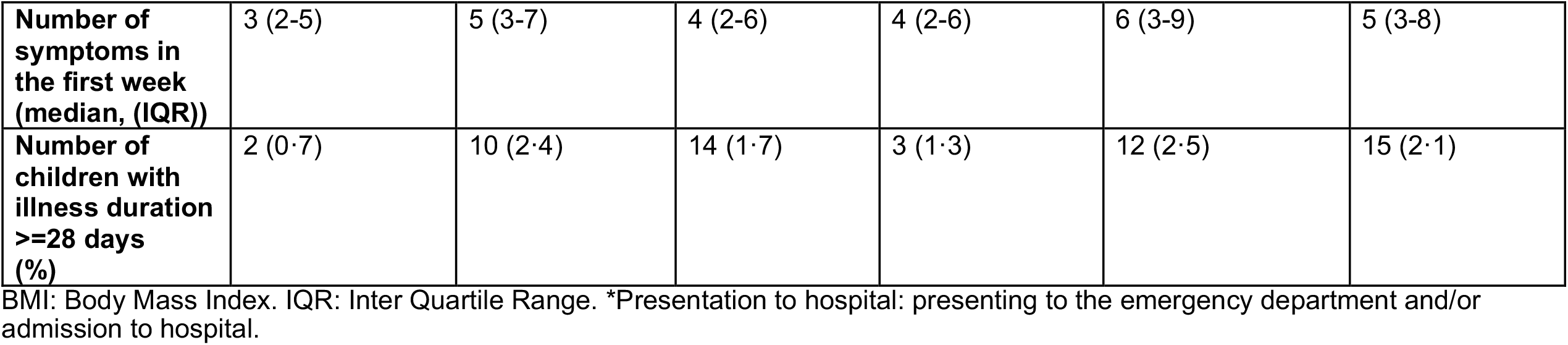
Characteristics of UK school-aged children presenting with COVID-19, during periods of SARS-CoV-2 Alpha or Delta variant predominance.

|  | Overall symptomatic children with positive SARS-CoV-2 test |  |  |  |  |  |
| --- | --- | --- | --- | --- | --- | --- |
|  | Alpha variant period |  |  | Delta variant period |  |  |
|  | Younger children<br>(aged 5-11 years) | Older children<br>(aged 12-17 years) | Overall cohort<br>(aged 5-17 years) | Younger children<br>(aged 5-11 years) | Older children<br>(aged 12-17 years) | Overall cohort<br>(aged 5-17 years) |
| **Number** | 276 | 418 | 694 | 227 | 479 | 706 |
| **Males (%)** | 144 (52.2) | 213 (50.9) | 357 (51.4) | 102 (44.9) | 255 (53.2) | 357 (50.6) |
| **Age, years**<br>**(median, (IQR))** | 9 (7-10) | 15 (13-16) | 12 (9-15) | 9 (8-11) | 15 (13-16) | 14 (13-16) |
| **BMI (kg/m<sup>2</sup>)**<br>**(median, (IQR))** | 16.44 (14.79-18.90) | 19.53 (17.24-22.07) | 18.16 (15.97 - 21.16) | 16.68 (14.61-19.27) | 18.97 (16.88-21.31) | 18.21 (16.01 - 20.96) |
| **Asthma (%)** | 32 (11.6) | 49 (11.7) | 81 (11.6) | 24 (10.6) | 56 (11.7) | 80 (11.3) |
| **Heart disease (%)** | 0 (0.0) | 3 (0.9) | 3 (0.4) | 2 (0.7) | 1 (0.2) | 3 (0.4) |
| **Diabetes (%)** | 1 (0.4) | 4 (1.0) | 5 (0.8) | 1 (0.4) | 1 (0.2) | 2 (0.3) |
| **Renal disease (%)** | 0 (0.0) | 0 (0.0) | 0 (0.0) | 0 (0.0) | 1 (0.2) | 4 (0.1) |
| **Number of children**<br>**presenting to hospital\* (%)** | 6 (2.2) | 8 (1.9) | 14 (2.0) | 8 (3.5) | 8 (1.7) | 16 (2.2) |
| **Illness duration**<br>**(median, (IQR))** | 4 (2-8) | 6 (3-10) | 5 (2-9.75) | 4 (2-7.5) | 6 (3-10) | 5 (2-9) |
| **Number of symptoms in the first week (median, (IQR))** | 3 (2-5) | 5 (3-7) | 4 (2-6) | 4 (2-6) | 6 (3-9) | 5 (3-8) |
| **Number of children with illness duration >=28 days (%)** | 2 (0·7) | 10 (2·4) | 14 (1·7) | 3 (1·3) | 12 (2·5) | 15 (2·1) |

BMI: Body Mass Index. IQR: Inter Quartile Range. \*Presentation to hospital: presenting to the emergency department and/or admission to hospital.

**Individual symptom duration** was short (median 5 days) (Figure 4). The symptom of longest duration was ‘blisters’ (particularly in younger children with Alpha infection); however, the prevalence of blisters was extremely low (approx.1%) (Supplementary Table 3).

**Figure 4.**
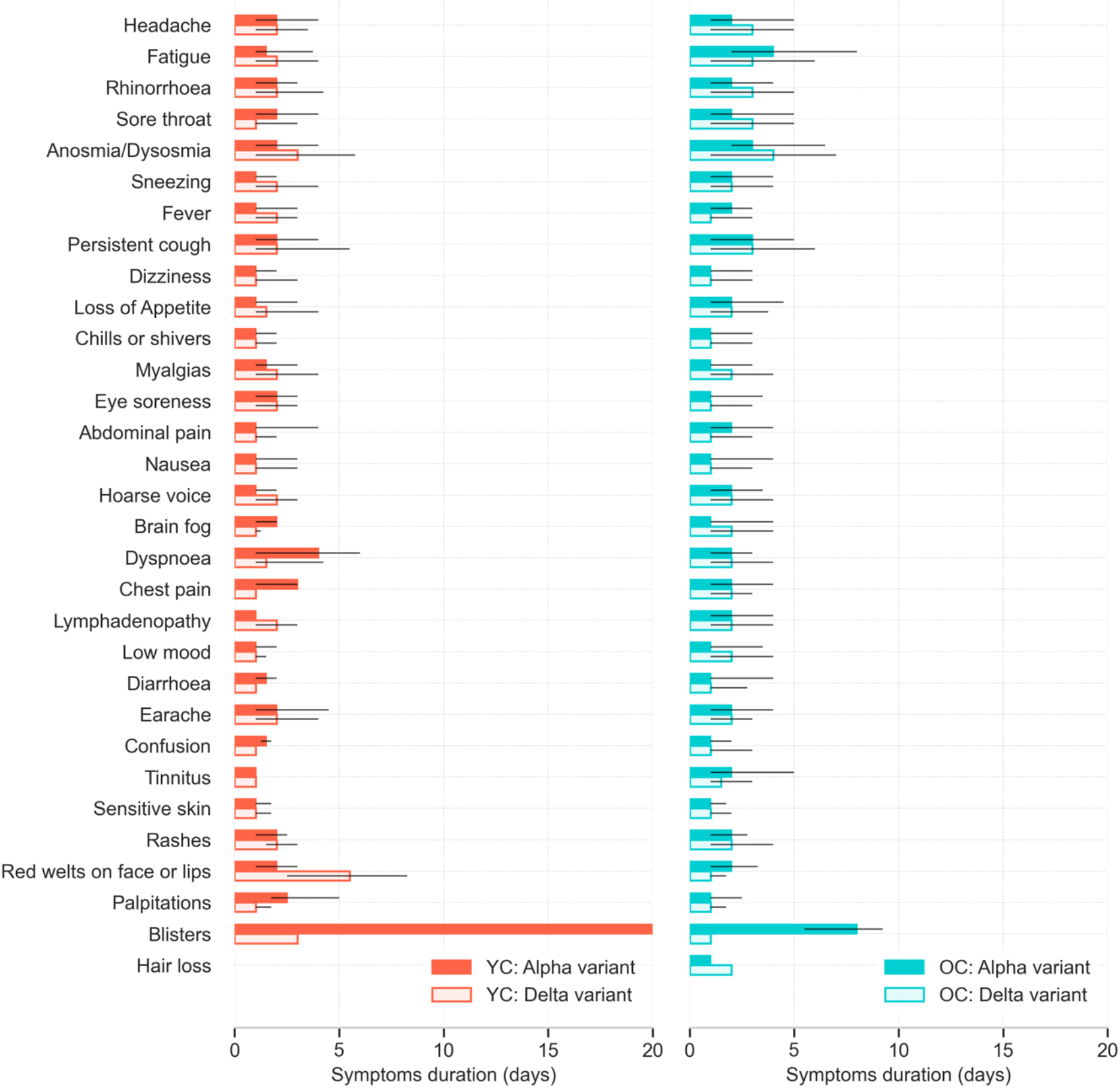
Median duration and IQR of each symptom reported over the course of illness in younger (5–11 years) and older (12–17 years) children with COVID-19, whose illness lasted <28 days, during periods of SARS-CoV-2 Alpha or Delta variant predominance.

**Presentation to hospital** was reported for 14 younger and 16 older children: 6 [2.2%] of 276 younger and 8 [1.9%] of 418 older children with Alpha infection and 8 [3.5%] of 227 younger and 8 [1·7%] of 479 older children with Delta infection (Table 1).

**Illness longer than 28 days** was reported for 5 younger and 22 older children overall (Table 1). Illness had resolved by 28 days in 274 [99.3%] of 276 younger, and in 408 [97.6%] of 418 older children with Alpha infection; and in 224 [98.7%] of 227 younger, and in 467 [97.5%] of 479 older children with Delta infection. For those children with longer illness duration, no new symptom developed after day 28.

Symptoms in children testing negative for SARS-CoV-2 during the same Alpha and Delta timeframes are presented in Supplementary Table 5 and Supplementary Figure 1. Rhinorrhoea, sore throat, sneezing, and persistent cough were more common during the Delta period, and headache and fatigue during the Alpha period (not statistically tested).

## Discussion

Here we present the illness profile of COVID-19 in UK school-aged children with SARS-CoV-2 Delta (B.1.617.2) variant infection, and provide similar data for COVID-19 due to the Alpha (B.1.1.7) variant. Several individual symptoms were more common with Delta infection (particularly in older children), and symptom burden appeared slightly greater, in both age groups. However, most symptoms were of short duration (median, <2 days); overall illness duration was short (median, 5 days); few children required hospital attention; and illness lasting for more than 28 days was uncommon. Our data suggest that clinical characteristics of COVID-19 due to the Delta variant in children are broadly similar to COVID-19 due to other variants.^13^

For both variants, the symptom with longest duration was ‘blisters’, although this symptom was rare. Five types of cutaneous lesions have been identified with COVID-19^17^ of which pseudo-chilblains, mainly reported in children, are at the lowest spectrum of severity; these resolve spontaneously, albeit slowly, in most cases. As children were not clinically examined, nor photos submitted, we are not able to comment further on the nature of these lesions.

To our knowledge, this is the first large-scale epidemiological study comparing illness characteristics of children with Alpha vs. Delta infection. The CLoCk matched-cohort study reported recently on a UK cohort tested from January to March 2021, which was during Alpha variant predominance (https://www.researchsquare.com/article/rs-798316/v1). The UK Office for National Statistics (ONS) has examined prevalence of prolonged symptoms in children at several timepoints across the pandemic but to date no comparison of symptoms between time points has been presented (https://www.ons.gov.uk/peoplepopulationandcommunity/healthandsocialcare/conditionsanddiseases/bulletins/prevalenceofongoingsymptomsfollowingcoronaviruscovid19infectionintheuk/2september2021). Whilst cohort studies have included some comparisons between variants in the adult- and child-populations considered jointly, child-specific data have not been reported separately.^9^ The latest report from the USA Centers for Disease Control indicates that differences in disease severity and duration between Delta and previous variants in children are unclear (https://www.cdc.gov/mmwr/volumes/70/wr/mm7036e1.htm?s_cid=mm7036e1_w). However, although USA cases in children have increased recently, the proportion of hospitalised children requiring intensive care is unchanged, suggesting infection severity is at least no worse (and possibly less) with evolution of circulating variants.^18^ In England^9^ and Scotland,^19^ observational data from the population as a whole (no child-specific aggregation) suggest hospitalization may be higher with Delta compared to Alpha infection (noting that vaccination has changed the profile of illness in adults, and currently the majority of hospitalized adults in the UK are unvaccinated). In contrast, data from Norway (again, children not analysed separately) did not support higher rates of hospitalisation with the Delta variant.^20^

With the exception of odds ratios for symptom presence, which could be robustly tested, our data are descriptive. We included data from all regularly proxy-logged children with a positive test during the two timeframes, noting the many differences between the two groups that could not be controlled for. As for many children world-wide, the pandemic has been extremely disruptive for UK schooling. Lockdown was re-imposed across the UK just before Christmas 2020; and primary and secondary schools were closed throughout January and February 2021. Schools reopened from March 8 until the end of the school year (nominally, end of July) (https://www.gov.uk/government/publications/covid-19-response-spring-2021). However, school attendance continued to be compromised on an *ad hoc* and intermittent basis for many children (for example, at-home quarantining whilst awaiting a test result, whether for the child personally, a family member, or classmates). Differences in school attendance may have affected our data in multiple ways, including differing daily routines (e.g., linking school attendance with proxy-reporting habits), exposure to SARS-CoV-2 at school and on public transport, and testing of children according to parental decisions regarding school attendance.

The UK national lockdown was progressively lifted after March 29, 2021, which altered prevalence of SARS-CoV-2 and other circulating respiratory viruses. Rhinovirus prevalence, which had increased rapidly at the start of the school year in September 2020, suddenly fell in January 2021, only to rise again from March 2021 (concordant with school re-opening) (https://assets.publishing.service.gov.uk/government/uploads/system/uploads/attachment_data/file/1016276/Weekly_Flu_and_COVID-19_report_w36.pdf figure 16). Other tested viruses (adenovirus, human metapneumovirus, respiratory syncytial virus and parainfluenza) were at unusually low levels during January to April 2021.

With lifting of lockdown, a rapid and unseasonal increase in parainfluenza prevalence was observed, followed by a rise of respiratory syncytial virus to unusually high levels (which continues high at time of writing). Thus, we considered whether our data might inadvertently capture symptoms from viruses other than COVID-19 - noting here that all children included in the Alpha and Delta COVID-19 groups had a positive SARS-CoV-2 test (i.e., co-infection). Although we could not formally test this, we have included illness profiles from symptomatic children testing negative for SARS-CoV-2 during the two timeframes; rhinorrhoea, sore throat, sneezing, and persistent cough were more common during the Delta period with other symptoms less common. Against co-infection, though, was the commonality of the top seven symptoms with either SARS-CoV-2 variant and other similarities in illness profiles, as detailed above. Without simultaneous testing for other circulating viruses the possibility of co-infection cannot be determined.

There were also large seasonal differences between the two timeframes (mid-winter to mid-Spring vs. early to mid-summer), and, associated with this, varying prevalence of seasonal allergic rhinitis.

Testing policy also changed over time. At the start of this study, access to SARS-CoV-2 testing was symptom-based (informed by data from adults),^14^ and required the presence of persistent cough, fever, and/or anosmia/dysosmia (https://www.nhs.uk/conditions/coronavirus-covid-19/symptoms/main-symptoms/). However, many other symptoms are manifest in infected individuals (both adults and children)^13,15,21^ with headache and fatigue the most common symptoms in children.^13^ Consistent with our previous work^13^ and that of others,^22^ the current study supports inclusion of other symptoms, particularly headache, if testing access in children is gated by symptom manifestation. Of note, regular (twice-weekly) rapid-testing of asymptomatic teachers and secondary school children was in place from school reopening.

We recognise other weaknesses of our study. Our study only assesses illness profile in symptomatic children. We are not able to comment on relative transmissibility or on prevalence of asymptomatic infection with Alpha vs. Delta variants. We only present data for symptoms assessed by direct questioning through the app across both timeframes (Supplementary Table 1). Any novel symptoms unique to Delta variant would not be captured by our study. Our current study does not present illness profiles in children with illness for more than 28 days, to avoid bias introduced by our census timing, although we do report the numbers of children with long illness duration which were low with either variant. Our previous study showed that prolonged illness duration in children with COVID-19 is uncommon^13^ consistent with the most recent (September 16, 2021) publication of the UK ONS Coronavirus Infection Survey: 3.3% (95%CI 2.5-4.5) of children aged 2-11 years testing positive for SARS-CoV-2 had one of 12 symptoms 29-56 days following infection, compared to 3.6% (95%CI 2.7-4.8) in a matched negative control group (noting that this latter study may be more representative of children in the general population than our own,^23^ and includes children with asymptomatic infection) (https://www.ons.gov.uk/peoplepopulationandcommunity/healthandsocialcare/conditionsanddiseases/articles/technicalarticleupdatedestimatesoftheprevalenceofpostacutesymptomsamongpeoplewithcoronaviruscovid19intheuk/26april2020to1august2021). Lastly, and crucially, proxy-reported children depended on an adult participating in this citizen science initiative, who was willing and able to report through a smart-phone app. Awareness of COVID-19 symptoms may also have changed over time in reporters, which may have influenced their proxy-reporting for children.

## Conclusions

In conclusion, our data show that COVID-19 due to Delta variant B.1.617.2 in UK school-aged children is usually of short duration and low symptom burden, similar to COVID-19 due to the Alpha variant B.1.1.7. and with the seven most prevalent symptoms common to both strains. Few proxy-reported children presented to hospital, and the numbers of children with illness >28 days were low. Our study adds quantitative information around manifestation of infection due to the Delta variant in children, which will contribute to the ongoing debate regarding the role of vaccination in children, particularly for younger children.

## Data Availability

Data collected in the ZOE COVID Study App can be shared with other health researchers through the UK National Health Service-funded Health Data Research UK and Secure Anonymised Information Linkage consortium, housed in the UK Secure Research Platform (Swansea, UK). Anonymised data are available to be shared with researchers according to their protocols in the public interest https://web.www.healthdatagateway.org/dataset/594cfe55-96e3-45ff-874c-2c0006eeb881.

https://arxiv.org/abs/2011.00867

## Abbreviations

CI: Confidence Interval
COVID-19: coronavirus disease 2019
FDR: False discovery rate
IQR: inter-quartile range
KCL: King’s College London
LFAT: Lateral flow antigen test
ONS: Office for National Statistics (UK)
PCR: polymerase chain reaction
SARS-CoV-2: severe acute respiratory syndrome-related coronavirus 2
UK: United Kingdom of Great Britain and Northern Ireland
USA: United States of America
WHO: World Health Organization

