## Supplementary Material for "Illness characteristics of COVID-19 in children infected with the SARS-CoV-2 Delta variant"

**Supplementary Methods.**

The COVID Symptom Study (administered through the ZOE COVID Study App) was launched jointly by ZOE Limited and King's College London on March 24, 2020. Data were acquired through a mobile application.[24] At time of registration through the app, all participants provide consent for their data to be used for research. Participants can withdraw from the study at any time, with their data, (including proxy-reported data) subsequently excluded from analysis.

The app was tested both before launch and before version updates. The app usage workflow was described in the earliest publication about the COVID Symptom Study (doi: 10.1126/science.abc0473). The team of app developers monitor the statistics of app usage and users' input daily, to assess for faults (e.g., problems in the app downloads or logging). No major disruptions were detected during this study.

Conjointly to the app development, software for data extraction, curation and analytics was developed, which guarantees the repeatability of the data curation over time and the consistency on different timestamps, according to stringent predefined criteria (filters). The software, based on Python language, is called ExeTera; documentation and associated code can be downloaded here: https://arxiv.org/abs/2011.00867

Currently 92% of UK adults own a smartphone, with little difference according to socioeconomic status (https://www.statista.com/statistics/300384/mobile-phone-usage-in-the-uk-by-socio-economic-group/).

**Supplementary Table 1. Incidence (%) of the Alpha and Delta variants of SARS-CoV-2 in United Kingdom,** in two timeframes: December 28, 2020 to May 6, 2021, and May 26, 2021 to July 8, 2021 (https://assets.publishing.service.gov.uk/government/uploads/system/uploads/attachment_data/file/975754/Variants_of_Concern_Technical_Briefing_8_Data_England.xlsx; GISAID https://covariants.org).

| Week of | Alpha (B.1.1.7) | Delta (B.1.617.2) |
|---|---|---|
| 30 December 2020 | 78·1 | 0 |
| 6 January | 82·3 | 0 |
| 13 January | 87·6 | 0 |
| 20 January | 90·8 | 0 |
| 27 January | 93·8 | 0 |
| 3 February | 95·7 | 0 |
| 10 February | 97·4 | 0 |
| 17 February | 98·1 | 0 |
| 24 February | 98·6 | 0 |
| 3 March | 98·8 | 0 |
| 10 March | 98·9 | 0 |
| 17 March | >99 | 0 |
| 24 March | 98 | 0 |
| 31 March | 98 | 0 |
| 7 April | 95 | 2 |
| 14 April | 92 | 6 |
| 21 April | 87 | 9 |
| 28 April | 76 | 24 |
| 5 May | 71·5 | 25·8 |
| 26 May | 19·3 | 80·0 |
| 2 June | 9·4 | 90·2 |
| 9 June | 5 | 95 |
| 16 June | 2 | 98 |
| 23 June | 2 | 98 |
| 30 June | <1 | >99 |
| 7 July | <1 | >99 |

**Supplementary Table 2. List of symptom questions asked by the ZOE COVID Study app during the current study period**.

| **Symptom** | **COVID Symptom Study app question** |
|---|---|
| Fever | Fever (at least 37.8C or 100F) |
| Persistent Cough | Persistent cough (coughing a lot for more than an hour or 3 or more coughing episodes in 24 hours) |
| Fatigue | Unusual fatigue (no; mild fatigue; severe fatigue/ I struggle to get out of bed) |
| Dyspnoea | Shortness of breath or trouble breathing (no; yes mild symptoms/ slight shortness of breath during ordinary activity: yes significant symptoms/ breathing is comfortable only at rest; yes, severe symptoms/ breathing is difficult even at rest). |
| Anosmia/Ageusia | Loss of smell / taste |
| Hoarse Voice | Unusually hoarse voice |
| Chest Pain | Unusual chest pain or tightness in your chest |
| Abdominal Pain | Unusual abdominal pain or stomach-ache |
| Diarrhoea | Diarrhoea |
| Stools | How many loose stools in the last 24 hours? |
| Confusion | Confusion, disorientation or drowsiness |
| Eye Soreness | Do your eyes have any unusual eye-soreness or discomfort (e.g. light sensitivity, excessive tears, or pink/red eye)? |

| | |
|---|---|
| Loss of appetite | Skipping meals |
| Headache | Headache |
| Nausea | Nausea or vomiting |
| Dizziness | Dizziness or light-headedness |
| Sore Throat | Sore or painful throat |
| Myalgias | Unusual strong muscle pains or aches |
| Red Welts | Raised, red, itchy welts on the skin or sudden swelling of the face or lips |
| Blisters | Red/purple sores or blisters on your feet, including your toes |
| Allergy Exacerbation | Increase in your usual allergy symptoms |
| Rashes | Rash on your arms or torso |
| Sensitive Skin | Strange, unpleasant sensations in your skin like pins & needles or burning |
| Hair Loss | Unusual hair loss |
| Low Mood | Feeling down, depressed, or hopeless |
| Brain Fog | Loss of concentration or memory (brain fog) |
| Dysosmia/Dysgeusia | Altered smell / taste (things smell or taste different to usual) |
| Rhinorrhoea | Runny nose |
| Sneezing | Sneezing more than usual |
| Ear Pain | Earache |

| | |
|---|---|
| Tinnitus | Ringing in your ears |
| Lymphadenopathy | Swollen neck glands |
| Palpitations | Unusually fast or irregular heartbeat (palpitations) |
| Arthralgias | Unusual joint pains or aches |
| Mouth Ulcers | Mouth or tongue ulcers |
| Tongue Changes | Changes to tongue surface |

**Supplementary Table 3. Symptoms over the entire illness duration in children testing positive for Alpha and Delta variants of SARS-CoV-2.** Data refers to younger (age 5–11 years) and older (age 12-17 years) children. Only children whose illness duration was <28 days are included. Symptom onset between December 28, 2020 and May 6, 2021 was attributed to infection with the Alpha variant, and symptom onset between May 26, 2021 and July 8, 2021 to infection with the Delta variant.

| | Younger children | | Older children | | All children | |
|---|---|---|---|---|---|---|
| | Alpha (n=276) | Delta (n=227) | Alpha (n=418) | Delta (n=479) | Alpha (n=694) | Delta (n=706) |
| Headache | 110 (39·9) | 138 (60·8) | 257 (61·5) | 353 (73·7) | 367 (52·9) | 491 (69·5) |
| Fatigue | 107 (38·8) | 111 (48·9) | 238 (56·9) | 288 (60·1) | 345 (49·7) | 399 (56·5) |
| Rhinorrhoea | 130 (47·1) | 122 (53·7) | 198 (47·4) | 276 (57·6) | 328 (47·3) | 398 (56·4) |
| Sore throat | 90 (32·6) | 83 (36·6) | 191 (45·7) | 290 (60·5) | 281 (40·5) | 373 (52·8) |
| Sneezing | 96 (34·8) | 82 (36·1) | 131 (31·3) | 167 (34·9) | 227 (32·7) | 249 (35·3) |
| Anosmia/Dysosmia | 41 (14·9) | 45 (19·8) | 167 (40·0) | 224 (46·8) | 208 (30·0) | 269 (38·1) |
| Fever | 88 (31·9) | 105 (46·3) | 113 (27·0) | 207 (43·2) | 201 (29·0) | 312 (44·2) |
| Persistent cough | 74 (26·8) | 60 (26·4) | 108 (25·8) | 148 (30·9) | 182 (26·2) | 208 (29·5) |
| Loss of appetite | 46 (16·7) | 43 (18·9) | 80 (19·1) | 122 (25·5) | 126 (18·2) | 165 (23·4) |
| Abdominal pain | 53 (19·2) | 48 (21·1) | 66 (15·8) | 65 (13·6) | 119 (17·1) | 113 (16·0) |
| Dizziness | 26 (9·4) | 25 (11·0) | 86 (20·6) | 153 (31·9) | 112 (16·1) | 178 (25·2) |
| Chills or shivers | 27 (9·8) | 41 (18·1) | 78 (18·7) | 114 (23·8) | 105 (15·1) | 155 (22·0) |
| Myalgias | 18 (6·5) | 14 (6·2) | 77 (18·4) | 80 (16·7) | 95 (13·7) | 94 (13·3) |
| Eye soreness | 19 (6·9) | 34 (15·0) | 74 (17·7) | 107 (22·3) | 93 (13·4) | 141 (20·0) |
| Nausea | 26 (9·4) | 30 (13·2) | 60 (14·4) | 80 (16·7) | 86 (12·4) | 110 (15·6) |
| Hoarse voice | 31 (11·2) | 37 (16·3) | 53 (12·7) | 98 (20·5) | 84 (12·1) | 135 (19·1) |
| Confusion | 9 (3·3) | 9 (4·0) | 42 (10·0) | 56 (11·7) | 51 (7·3) | 65 (9·2) |

| | | | | | | |
|---|---|---|---|---|---|---|
| Diarrhoea | 20 (7·2) | 18 (7·9) | 27 (6·5) | 36 (7·5) | 47 (6·8) | 54 (7·6) |
| Dyspnoea | 9 (3·3) | 6 (2·6) | 37 (8·9) | 50 (10·4) | 46 (6·6) | 56 (7.9) |
| Low mood | 15 (5·4) | 7 (3·1) | 29 (6·9) | 33 (6·9) | 44 (6·3) | 40 (5·7) |
| Chest pain | 11 (4·0) | 7 (3·1) | 32 (7·7) | 43 (9·0) | 43 (6·2) | 50 (7·1) |
| Lymphadenopathy | 12 (4·3) | 12 (5·3) | 30 (7·2) | 38 (7·9) | 42 (6·1) | 50 (7·1) |
| Earache | 11 (4·0) | 14 (6·2) | 24 (5·7) | 41 (8·6) | 35 (5·0) | 55 (7·8) |
| Rash | 15 (5·4) | 9 (4·0) | 13 (3·1) | 19 (4·0) | 28 (4·0) | 28 (4·0) |
| Delirium | 2 (0·7) | 5 (2·2) | 23 (5·5) | 38 (7·9) | 25 (3·6) | 43 (6·1) |
| Tinnitus | 6 (2·2) | 7 (3·1) | 16 (3·8) | 21 (4·4) | 22 (3·2) | 28 (4·0) |
| Sensitive skin | 6 (2·2) | 7 (3·1) | 14 (3·3) | 22 (4·6) | 20 (2·9) | 29 (4·1) |
| Red welts on face or lips | 8 (2·9) | 5 (2·2) | 9 (2·2) | 10 (2·1) | 17 (2·4) | 15 (2·1) |
| Palpitations | 4 (1·4) | 6 (2·6) | 10 (2·4) | 7 (1·5) | 14 (2·0) | 13 (1·8) |
| Blisters | 1 (0·4) | 1 (0·4) | 5 (1·2) | 5 (1·0) | 6 (0·9) | 6 (0·8) |
| Hair loss | 0 (0·0) | 0 (0·0) | 1 (0·2) | 2 (0·4) | 1 (0·1) | 2 (0·3) |

**Supplementary Table 4. Odds ratios for symptoms presenting with SARS-CoV-2 infection, comparing Delta to Alpha infection.** Odds ratios are adjusted for age and sex. Only children with illness duration <28 days are included. The bold font and asterisk indicate statistical significance after FDR correction with α>0.05. Symptoms are ranked based on prevalence in the overall cohort during the period of Alpha predominance.

| | Younger children | | Older children | | All children | |
|---|---|---|---|---|---|---|
| | Odd Ratio (CI) | p-value | Odd Ratio (CI) | p-value | Odd Ratio (CI) | p-value |
| Headache | **2·31 (1·60-3·33)** | **<0·001*** | **1·81 (1·36-2·42)** | **<0·001*** | **1·97 (1·57-2·46)** | **<0·001*** |
| Fatigue | 1·46 (1·02-2·09) | 0·039 | 1·16 (0·89-1·52) | 0·280 | 1·25 (1·01-1·55) | 0·038 |
| Rhinorrhoea | 1·31 (0·92-1·86) | 0·139 | **1·52 (1·17-1·98)** | **0·002*** | **1·44 (1·17-1·78)** | **0·001*** |
| Sore throat | 1·13 (0·77-1·64) | 0·534 | **1·87 (1·43-2·45)** | **<0·001*** | **1·58 (1·27-1·97)** | **<0·001*** |
| Sneezing | 1·09 (0·75-1·57) | 0·662 | 1·17 (0·88-1·54) | 0·278 | 1·14 (0·91-1·42) | 0·255 |
| Anosmia/Dysosmia | 1·29 (0·80-2·09) | 0·292 | 1·36 (1·04-1·78) | 0·025 | **1·36 (1·08-1·72)** | **0·010*** |
| Fever | **1·89 (1·31-2·73)** | **0·001*** | **2·05 (1·55-2·72)** | **<0·001*** | **1·99 (1·59-2·49)** | **<0·001*** |
| Persistent cough | 1·02 (0·68-1·51) | 0·942 | 1·29 (0·96-1·72) | 0·093 | 1·17 (0·92-1·48) | 0·193 |
| Loss of appetite | 1·18 (0·74-1·87) | 0·486 | 1·46 (1·06-2·01) | 0·02 | 1·35 (1·04-1·75) | 0·024 |
| Abdominal pain | 1·16 (0·75-1·81) | 0·503 | 0·83 (0·57-1·21) | 0·342 | 0·96 (0·72-1·27) | 0·752 |
| Dizziness | 1·12 (0·62-2·02) | 0·717 | **1·86 (1·37-2·54)** | **<0·001*** | **1·69 (1·29-2·21)** | **<0·001*** |
| Chills or shivers | 1·97 (1·17-3·33) | 0·011 | 1·38 (1-1·91) | 0·053 | **1·53 (1·16-2·01)** | **0·003*** |
| Myalgias | 0·91 (0·44-1·87) | 0·788 | 0·9 (0·64-1·27) | 0·554 | 0·91 (0·66-1·24) | 0·540 |
| Eye soreness | **2·40 (1·32-4·38)** | **0·004*** | 1·34 (0·97-1·87) | 0·080 | **1·55 (1·16-2·07)** | **0·003*** |
| Nausea | 1·43 (0·81-2·50) | 0·218 | 1·21 (0·84-1·74) | 0·315 | 1·27 (0·94-1·73) | 0·122 |
| Hoarse voice | 1·58 (0·94-2·65) | 0·083 | **1·78 (1·24-2·57)** | **0·002*** | **1·70 (1·26-2·28)** | **<0·001*** |
| Confusion | 1·17 (0·45-3·01) | 0·749 | 1·2 (0·79-1·84) | 0·396 | 1·21 (0·82-1·78) | 0·328 |
| Diarrhoea | 1·21 (0·62-2·37) | 0·578 | 1·16 (0·69-1·95) | 0·567 | 1·17 (0·78-1·76) | 0·451 |
| Dyspnoea | 0·77 (0·27-2·22) | 0·632 | 1·23 (0·79-1·93) | 0·366 | 1·15 (0·76-1·73) | 0·513 |
| Low mood | 0·52 (0·21-1·32) | 0·170 | 1·01 (0·6-1·7) | 0·972 | 0·85 (0·55-1·33) | 0·484 |
| Chest pain | 0·77 (0·29-2·04) | 0·604 | 1·22 (0·75-1·97) | 0·421 | 1·10 (0·72-1·68) | 0·670 |
| Lymphadenopathy | 1·22 (0·53-2·78) | 0·640 | 1·13 (0·69-1·86) | 0·635 | 1·15 (0·75-1·76) | 0·525 |
| Earache | 1·58 (0·70-3·58) | 0·267 | 1·58 (0·94-2·67) | 0·087 | 1·56 (1·00-2·42) | 0·048 |

| | | | | | | |
|---|---|---|---|---|---|---|
| Rash | 0·67 (0·29-1·58) | 0·361 | 1·29 (0·63-2·65) | 0·485 | 0·98 (0·58-1·68) | 0·951 |
| Delirium | 2·94 (0·56-15·38) | 0·202 | 1·48 (0·87-2·53) | 0·153 | 1·64 (0·98-2·72) | 0·058 |
| Tinnitus | 1·44 (0·47-4·43) | 0·529 | 1·16 (0·59-2·26) | 0·667 | 1·23 (0·70-2·17) | 0·477 |
| Sensitive skin | 1·39 (0·46-4·22) | 0·560 | 1·38 (0·7-2·74) | 0·352 | 1·41 (0·79-2·52) | 0·244 |
| Palpitations | 1·94 (0·54-7·01) | 0·313 | 0·62 (0·23-1·64) | 0·333 | 0·89 (0·42-1·92) | 0·773 |
| Red welts on face or lips | 0·75 (0·24-2·35) | 0·625 | 0·98 (0·39-2·43) | 0·962 | 0·88 (0·43-1·78) | 0·717 |
| Blisters | 1·11 (0·07-18·16) | 0·942 | 0·88 (0·25-3·06) | 0·838 | 0·92 (0·29-2·87) | 0·886 |
| Hair loss | 0·00 (0·00-0·00) | - | 2·15 (0·19-24·29) | 0·536 | 2·15 (0·19-24·29) | 0·535 |

*Statistically significant after FDR correction for multiple tests at 0·05

**Supplementary Table 5. Symptom presentation over the first 28 days of the disease in children testing negative for SARS-CoV-2.** Data refers to younger (age 5–11 years) and older (age 12-17 years) children, and the cohort overall. Individuals were grouped by symptom onset date, analogous to the positively-tested children (Alpha: between December 28, 2020 and May 6, 2021; Delta: between May 26, 2021 and July 8, 2021).

| | Younger children | | Older children | | All children | |
|---|---|---|---|---|---|---|
| | Alpha (n=3,748) | Delta (n=1,117) | Alpha (n=2,285) | Delta (n=231) | Alpha (n=6,033) | Delta (n=1,348) |
| Headache | 1,025 (27.3) | 255 (22·8) | 1,026 (44·9) | 98 (42·4) | 2,051 (34·0) | 353 (26·2) |
| Fatigue | 937 (25·0) | 189 (16·9) | 751 (32·9) | 63 (27·3) | 1,688 (28·0) | 252 (18·7) |
| Rhinorrhoea | 1,902 (50·7) | 673 (60·3) | 596 (26·1) | 123 (53·2) | 2,498 (41·4) | 796 (59·1) |
| Sore throat | 1,496 (39·9) | 497 (44·5) | 730 (31·9) | 130 (56·3) | 2,226 (36·9) | 627 (46·5) |
| Sneezing | 1,210 (32·3) | 391 (35·0) | 415 (18·2) | 89 (38·5) | 1,625 (26·9) | 480 (35·6) |
| Anosmia/Dysosmia | 188 (5·0) | 33 (3·0) | 160 (7·0) | 16 (6·9) | 348 (5·8) | 49 (3·6) |
| Fever | 1,212 (32·3) | 302 (27·0) | 484 (21·2) | 48 (20·8) | 1,696 (28·1) | 350 (26·0) |
| Persistent cough | 932 (24·9) | 422 (37·8) | 224 (9·8) | 63 (27·3) | 1,156 (19·2) | 485 (36·0) |
| Loss of appetite | 424 (11·3) | 97 (8·7) | 303 (13·3) | 16 (6·9) | 727 (12·1) | 113 (8·4) |
| Abdominal pain | 753 (20·1) | 142 (12·7) | 485 (21·2) | 30 (13·0) | 1,238 (20·5) | 172 (12·8) |
| Dizziness | 193 (5·1) | 32 (2·9) | 344 (15·1) | 32 (13·9) | 537 (8·9) | 64 (4·7) |
| Chills or shivers | 432 (11·5) | 89 (8·0) | 331 (14·5) | 20 (8·7) | 763 (12·6) | 109 (8·1) |
| Myalgias | 132 (3·5) | 23 (2·1) | 167 (7·3) | 12 (5·2) | 299 (5·0) | 35 (2·6) |
| Eye soreness | 215 (5·7) | 77 (6·9) | 209 (9·1) | 22 (9·5) | 424 (7·0) | 99 (7·3) |
| Nausea | 484 (12·9) | 106 (9·5) | 511 (22·4) | 28 (12·1) | 995 (16·5) | 134 (9·9) |
| Hoarse voice | 452 (12·1) | 166 (14·9) | 139 (6·1) | 30 (13·0) | 591 (9·8) | 196 (14·5) |
| Confusion | 63 (1·7) | 6 (0·5) | 129 (5·6) | 9 (3·9) | 192 (3·2) | 15 (1·1) |
| Diarrhoea | 258 (6·9) | 62 (5·6) | 210 (9·2) | 13 (5·6) | 468 (7·8) | 75 (5·6) |
| Dyspnoea | 101 (2·7) | 39 (3·5) | 99 (4·3) | 10 (4·3) | 200 (3·3) | 49 (3·6) |
| Low mood | 104 (2·8) | 11 (1·0) | 190 (8·3) | 7 (3·0) | 294 (4·9) | 18 (1·3) |

| | | | | | | |
|---|---|---|---|---|---|---|
| Chest pain | 71 (1·9) | 19 (1·7) | 114 (5·0) | 12 (5·2) | 185 (3·1) | 31 (2·3) |
| Lymphadenopathy | 236 (6·3) | 38 (3·4) | 159 (7·0) | 21 (9·1) | 395 (6·5) | 59 (4·4) |
| Earache | 154 (4·1) | 50 (4·5) | 132 (5·8) | 14 (6·1) | 286 (4·7) | 64 (4·7) |
| Rash | 148 (3·9) | 23 (2·1) | 97 (4·2) | 3 (1·3) | 245 (4·1) | 26 (1·9) |
| Delirium | 48 (1·3) | 13 (1·2) | 70 (3·1) | 3 (1·3) | 118 (2·0) | 16 (1·2) |
| Tinnitus | 38 (1·0) | 4 (0·4) | 64 (2·8) | 4 (1·7) | 102 (1·7) | 8 (0·6) |
| Sensitive skin | 38 (1·0) | 4 (0·4) | 64 (2·8) | 1 (0·4) | 102 (1·7) | 5 (0·4) |
| Palpitations | 18 (0·5) | 4 (0·4) | 45 (2·0) | 1 (0·4) | 63 (1·0) | 5 (0·4) |
| Red welts on face or lips | 89 (2·4) | 11 (1·0) | 58 (2·5) | 5 (2·2) | 147 (2·4) | 16 (1·2) |
| Blisters | 48 (1·3) | 2 (0·2) | 106 (4·6) | 0 (0·0) | 154 (2·6) | 2 (0·1) |
| Hair loss | 0 (0·0) | 0 (0·0) | 7 (0·3) | 1 (0·4) | 7 (0·1) | 1 (0·1) |

**Supplementary Figure 1. Prevalence of symptoms reported by negative COVID-19 testing in younger (aged 5–11 years) and older (aged 12–17 years) children, for symptoms presenting within 28 days, during periods of Alpha and Delta SARS-CoV-2 variant predominance.**

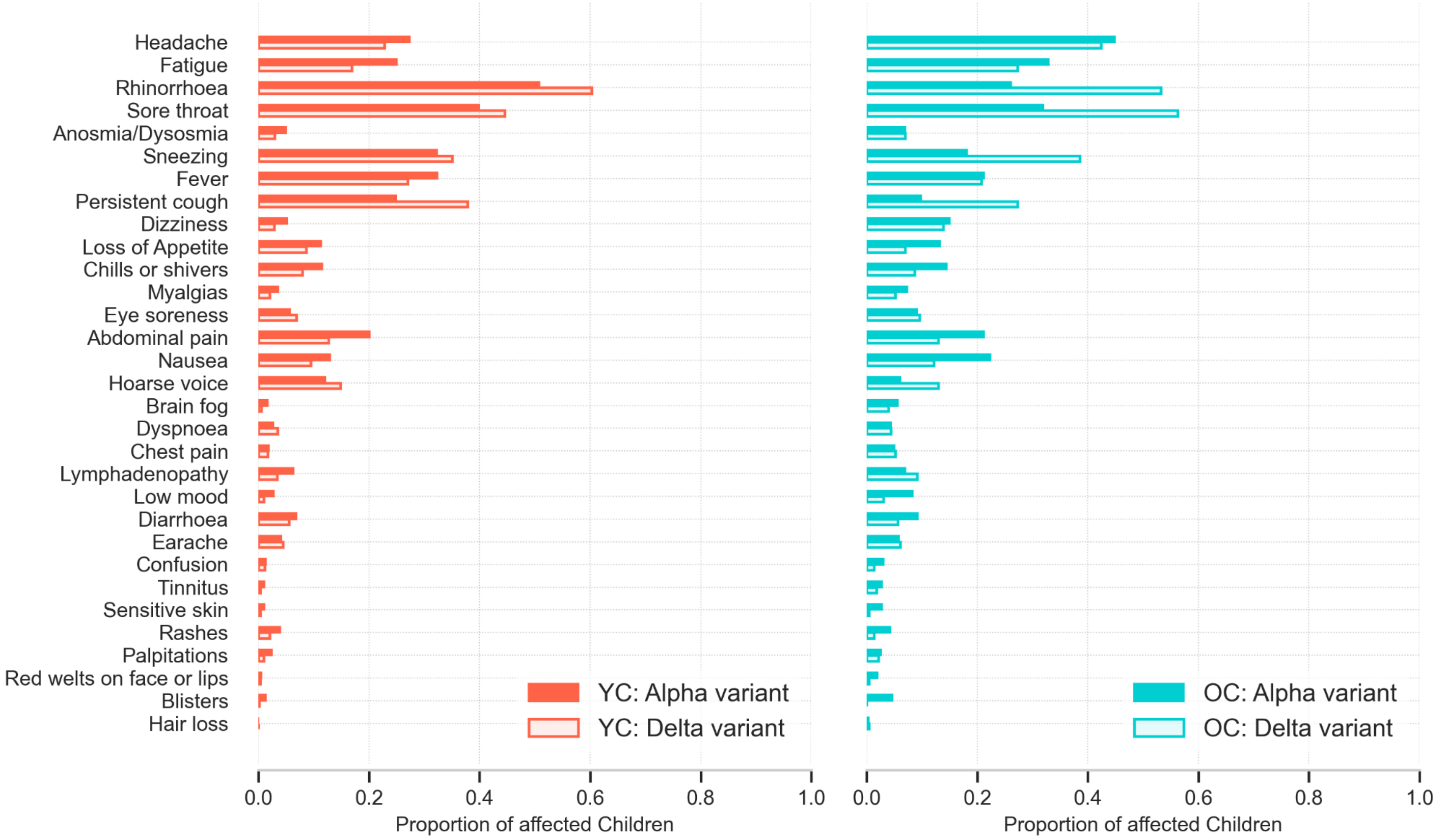
